# Using pet insurance claims to predict occurrence of vector-borne and zoonotic disease in humans

**DOI:** 10.1101/2024.08.09.24311752

**Authors:** Janice O’Brien, Aliya McCullough, Christian Debes, Audrey Ruple

**Affiliations:** Department of Population Health Sciences, Virginia-Maryland College of Veterinary Medicine, Virginia Tech, Blacksburg, VA, USA; Fetch Insurance Services, LLC, New York, NY, USA; Spryfox GmbH, Darmstadt, Germany

## Abstract

Taking a One Health approach to infectious diseases common to both dogs and people, pet insurance claims from 2008-2022 in the United States were compared to publicly available CDC-based data on human cases for Lyme disease, giardia, and Valley Fever (coccidioidomycosis). Despite having very different causative agents and etiologies, the disease trends for these three diseases were very similar between people and dogs both geographically and temporally. We furthermore demonstrated that adding dog data to the human data improves prediction models for those same diseases. With machine learning prediction tools for the pet insurance to increase prediction times and alert public health officials, pet insurance data could be a helpful tool to predict and detect diseases by estimating even earlier the effects of these common exposure diseases on human health. We also show the spatiotemporal distribution of intestinal worm diagnoses in dogs, and while it could not be directly compared to human data because the corresponding disease in humans (soil-transmitted helminths) has not been well monitored recently. However, these data can help inform researchers and public health workers.

## Introduction

In today’s highly interconnected world where human, animal, and environmental health are deeply related, novel disease detection methods are crucial. Identifying emerging and reemerging diseases at their earliest stages enables earlier public health interventions and therefore impacts more lives. Additionally, monitoring and modeling diseases provides insights into the epidemiology of these diseases, and doing so in multiple species helps to reveal the dynamics of the diseases in these different populations. The animal in the United States that shares the most environmental exposures with humans is arguably pet dogs.

Dogs have been explored as a sentinel of human disease across many different disease types: vector-borne pathogens,^1^ toxin exposures,^2^ and cancer.^3^ This is well covered by Sexton in the exploration of the shared exposome of people and dogs.^4^ Here we examined them as a potential sentinel for four infectious diseases with different types of pathogens and different etiologies: Lyme disease, giardiasis, Valley Fever (coccidioidomycosis), and soil-transmitted helminths.

Lyme disease is caused by the *Borrelia burgdorferi* bacterium transmitted by Ixodes ticks. It was first described in 1977 in Connecticut, but was present in ticks collected in Long Island in the 1940s^5^ and in Massachusetts white-footed mice collected in 1894,^6^ and so has been plaguing the northeastern United States for centuries. Since its recognition and development of appropriate testing methods in both dogs and people, Lyme disease has been apparently spreading geographically within the United States due to the spread of the Ixodes tick vector into new areas.^7^ The Ixodes spread has been shown to be secondary to factors like global climate change,^8^ spread through bird migration patterns,^9^ and deforestation and reforestation patterns.^10^ In humans, Lyme disease is frequently mis-diagnosed. In one study by Ferrouillet, only 40% of human general practitioners could accurately diagnose an erythema migrans rash^11^, which is considered pathognomonic for the disease. In another study by Gasmi,^12^ the misdiagnosis of erythema migrans rashes was 62.8%. This frequent misdiagnosis leads to people living with the disease for prolonged periods causing severe morbidity, and can even lead to the potentially lethal condition of Lyme carditis.^13^ In dogs, Lyme typically causes polyarthritis, fatigue, and loss of appetite. Untreated Lyme disease in dogs can lead to Lyme nephritis and death. The disease is zoonotic, but it cannot be shared directly between people and dogs – bites from infected ticks spread the disease. However, dogs and people living in the same areas share exposure to these ticks.

Giardiasis is a diarrheal disease caused by a protozoan parasite transmitted by the fecal-oral route, particularly via water contaminated with infected feces. Giardiasis in people and dogs is typically caused by different assemblages of the organism. Humans are usually infected with A and B, while dogs typically contract C and D, although both species can contract each other’s assemblages so zoonotic infection can occur.^14^ In both species, it is most frequently diagnosed in the young – children and puppies – and can be both severe and asymptomatic, depending on the individual infected. Both people and dogs that are otherwise compromised, such as those with weakened immune systems or the very young and very old, are more at risk for severe complications and death from the disease.

Valley Fever (coccidioidomycosis) is caused by inhaling spores of the coccidioides fungus, which typically grows in the soil. Dogs and people are infected by the same agent, and the disease course is very similar between the two species with primary infection occurring in the lungs, which can spread to other areas of the body. The infectious form is only found outside the body, so the disease is not contagious between individuals of either species, except in very rare cases. In people, the disease is often mis-diagnosed as a viral or bacterial infection, or even metastatic cancer,^15^ causing significant morbidity for those with delayed diagnoses. Previous studies have studied dogs as a possible sentinel of this agent when it moves into new geographic areas, as it has been proposed that dogs with digging and dirt-sniffing behaviors are more exposed to the disease than humans typically are. In Washington, cases were first identified in dogs before they were identified in people.^16^ In Texas, a study of 6087 of dogs used serological testing to identify geographic areas of exposure.^17^ A study conducted at the Veterinary Medical Teaching Hospital at University of California, Davis compared the geographic distribution of exposures among both dogs and people in California which were nearly identical.^18^ Despite all these previous studies proposing dogs as a sentinel for coccidioidomycosis, none have shown a direct comparison between human and canine incidence rates over time.

Soil-transmitted helminths are considered a neglected parasitic disease, and are the most common neglected tropical disease globally. Because of the morbidity caused by these infections, they have the capability to perpetuate poverty.^19^ Soil-transmitted helminth infections are thought to be rare in the USA, however recent work has demonstrated that there are likely persisting pockets of infection.^20,21^ The worms which cause disease in humans can come from many animals: wild canids, both feral and domestic cats, racoons, pigs, but domestic dogs are hosts to many of these parasites (*Ancylostoma caninum, A. brazilense, A. ceylanicum, Uncinaria stenocephala*, and *Toxocara canis*). When infected dogs defecate and those feces are left in the environment, contact or ingestion of the infective eggs or larvae can cause these infections in humans. The symptoms people experience are due to the bodily location in which the worms are migrating – through the skin, eyes, abdomen, etc. – and these migrations can cause significant health problems for the infected persons.

Dogs have previously been demonstrated as a sentinel of disease for people for several infectious diseases, so using pet insurance information to monitor the rates of these diseases in dogs is a reasonable step in a One Health approach to preventing and managing these diseases. Examining pet insurance data for disease insights and outcomes is beneficial because pet owners with pet insurance are more likely to opt for veterinary recommended diagnostics and treatment^22,23^ and less likely to elect economic euthanasia.^24,25^ Therefore, the clinical picture of disease in the pet is more likely to be complete.

## Results

### Pet population and diagnosis data

1,254,522.5 dog-years were represented in this population of insured dogs between 2008-2022, with 1,743,709 unique dogs. Among those dogs, there were a total of 36,419 diagnoses in the infectious conditions of tickborne disease (21%), giardia (37%), fungal disease (15%), and intestinal parasites (26%).

Within tickborne diseases, the diagnosis codes were: Lyme disease (3856, 49%), Babesiosis (37, 0.5%), Ehrlichiosis (763, 10%), Anaplasmosis (1367, 18%), and tick bite (1779, 23%). Within the intestinal parasites category, the top four diagnosis codes represented were: internal parasites (2885, 30%), coccidiosis (1560, 16%), hookworm (1324, 14%), and tapeworm (1286, 13%). Within the fungal disease group, the three diagnosis codes represented were: Valley Fever (Coccidioidomycosis) (4413, 80%), aspergillosis (241, 4%), and fungal infection (885, 16%).

The geographic distribution of the pet insurance dataset is generally representative of the US population density (see **Figure 1**). The age distribution of the insured population (see **Table 1**) was more heavily weighted toward younger dogs (the puppy and young adult age groups are the largest groups). There were more owner-reported purebred dogs compared to mixed breed dogs (63% to 36%).

**Figure 1:**
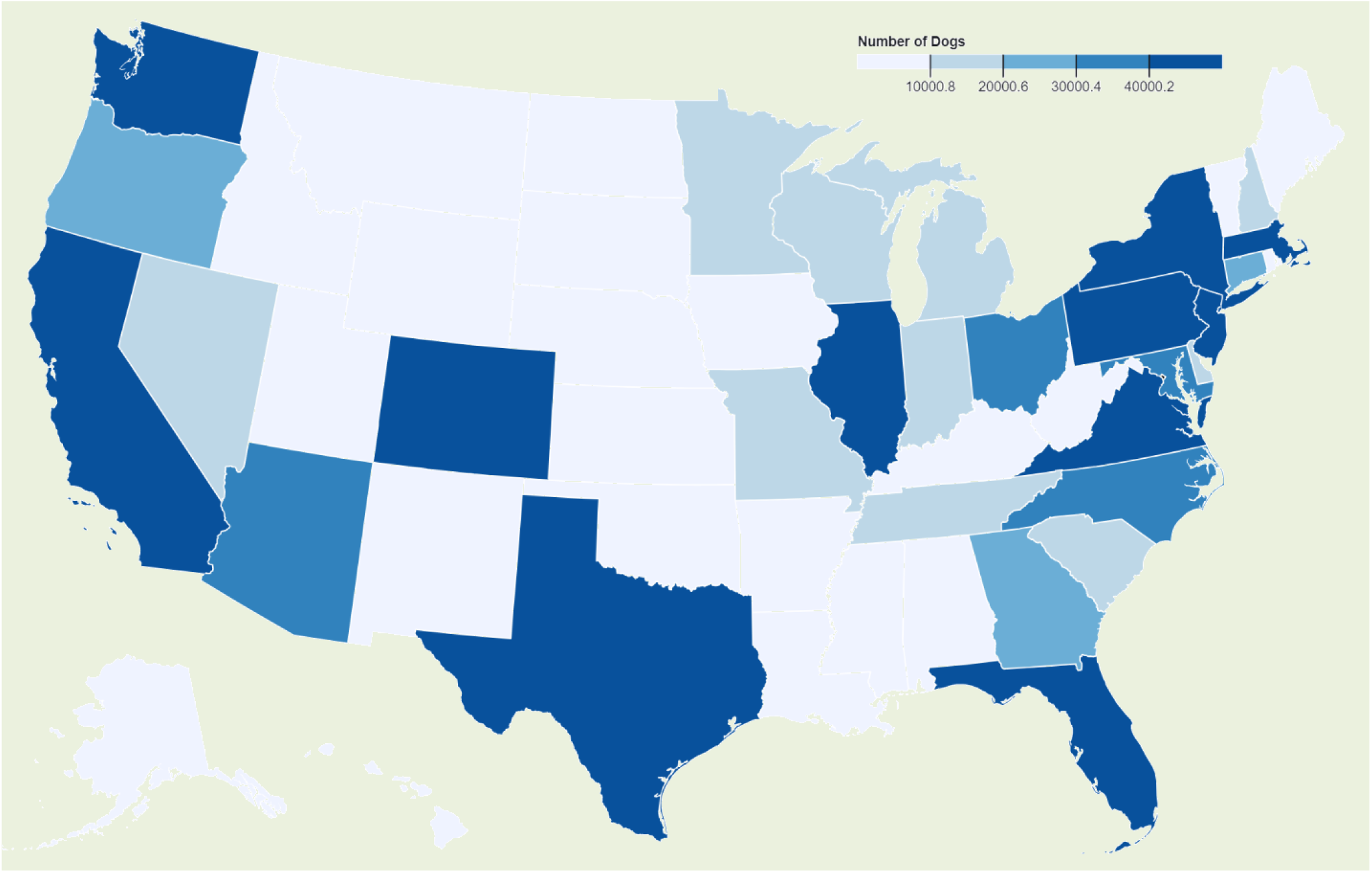
Geographic distribution of the dogs in this insurance dataset – counts

**Table 1:** Distribution of dogs by life stage and breed status across the disease categories.

| Category |  | Dogs diagnosed with Tickborne (7802) | Dogs diagnosed with Giardia (13489) | Dogs diagnosed with Fungal (5539) | Dogs diagnosed with Intestinal Parasites (9589) | Total Population (1743709) |
| --- | --- | --- | --- | --- | --- | --- |
| Life stage | Puppy | 26% (2025) | 83% (11204) | 12% (646) | 68% (6525) | 42% (382853) |
|  | Young Adult | 43% (3336) | 12% (1571) | 46% (2548) | 21% (2045) | 37% (338087) |
|  | Mature Adult | 22% (1723) | 4% (485) | 31% (1694) | 7% (699) | 15% (133982) |
|  | Geriatric | 9% (718) | 2% (229) | 12% (651) | 3% (320) | 6% (54240) |
| Breed | Purebred | 60% (4715) | 60% (8048) | 65% (3594) | 57% (5469) | 63% (1105223) |
|  | Mixed | 40% (3087) | 40% (5441) | 35% (1945) | 43% (4120) | 36% (638481) |

### Pet demographics across disease categories

Examining the changes in the affected population across the disease categories reveals both anticipated and unexpected findings, as shown in table 1. Puppies were overwhelmingly the largest group diagnosed with Giardia and intestinal parasites. Young adult dogs were the largest group diagnosed with tickborne and fungal diseases. There does not appear to be a difference between pure- or mixed-breed status and diagnoses with any of the diseases. There were some small differences between diseases, but in general the percentages of purebred were approximately 60% and mixed breed were approximately 40%.

### Comparison to Human disease trends

Here within each overall disease category, we highlight a specific disease of interest.

#### Lyme

The overall incidence rate of Lyme disease in dogs is approximately 6 times higher than that of humans. Dogs have a much longer season for the disease – starting in March and ending in September – while people mostly appear to have diagnoses starting in May. The geographic distribution of Lyme cases is nearly identical for the northeastern states and Minnesota/Wisconsin, but notably the incidence rate among dogs is higher in adjacent states like South Dakota, Illinois, Ohio, Michigan. The human and canine Lyme disease rates appear to follow the same general trends (looking at both national rates and for the top-15 states), with one important difference: that the canine rate changes appear to precede the changes in humans by approximately 1-2 years (see **Figure 2**).

**Figure 2:**
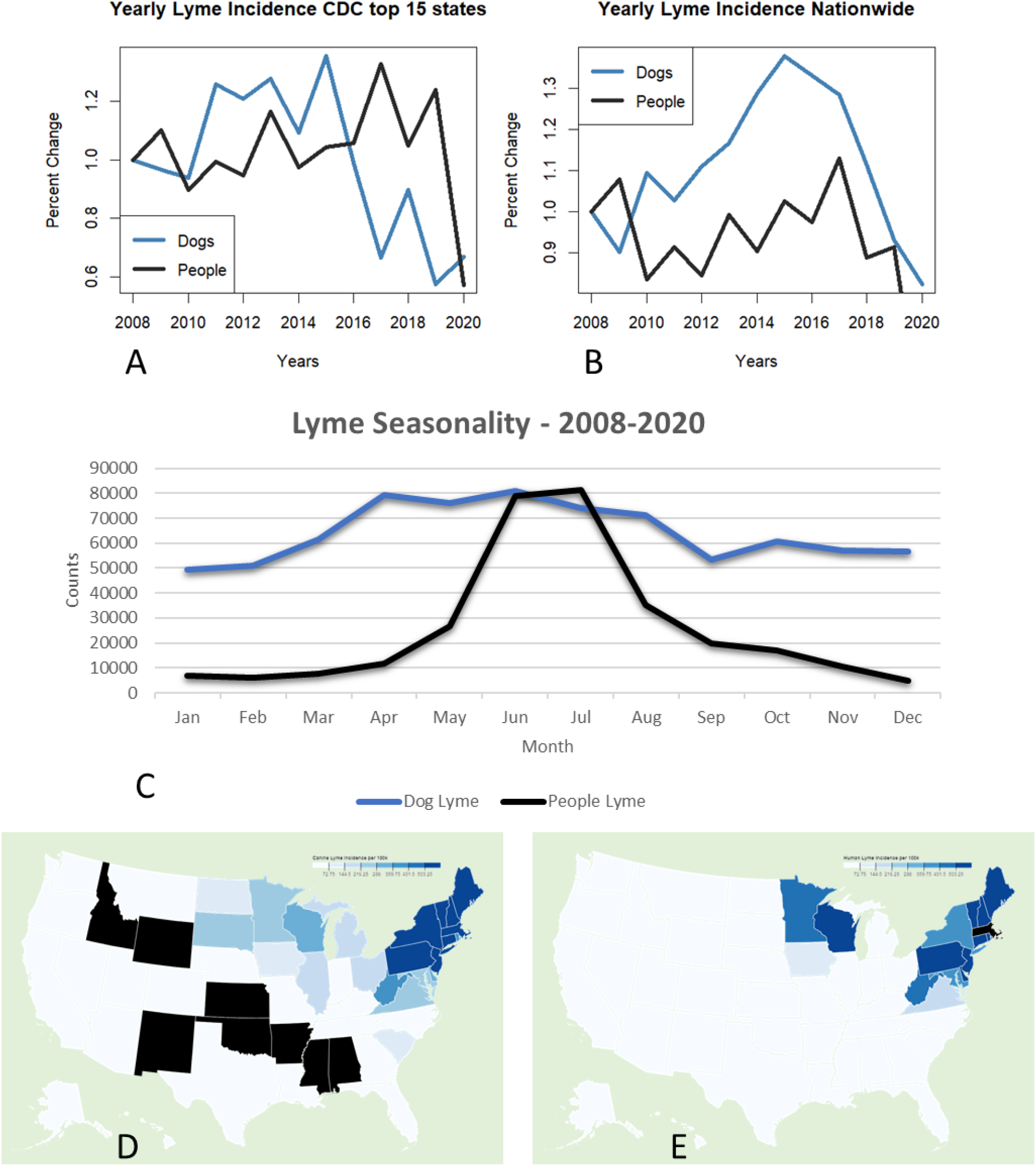
Yearly Lyme incidence rates for both humans and dogs for A) CDC top-15 Lyme states B) Nationally graphed relative to their incidence in 2008 C) Seasonal distribution of diagnosed Lyme case counts – 2008-2020 data overlayed. Canine cases multiplied by 200 so graphs can be compared relatively D) dog and E) human geographic distribution incidence per 100k in the United States

#### Giardia

Human and Canine Giardia incidence trends appear to follow a similar pattern, where canine cases peak in 2009, 2011, and 2015, and human cases peak in 2010 and 2016. Both species experienced lower relative incidence rates in 2013 and 2017 (see **Figure 3**).

**Figure 3:**
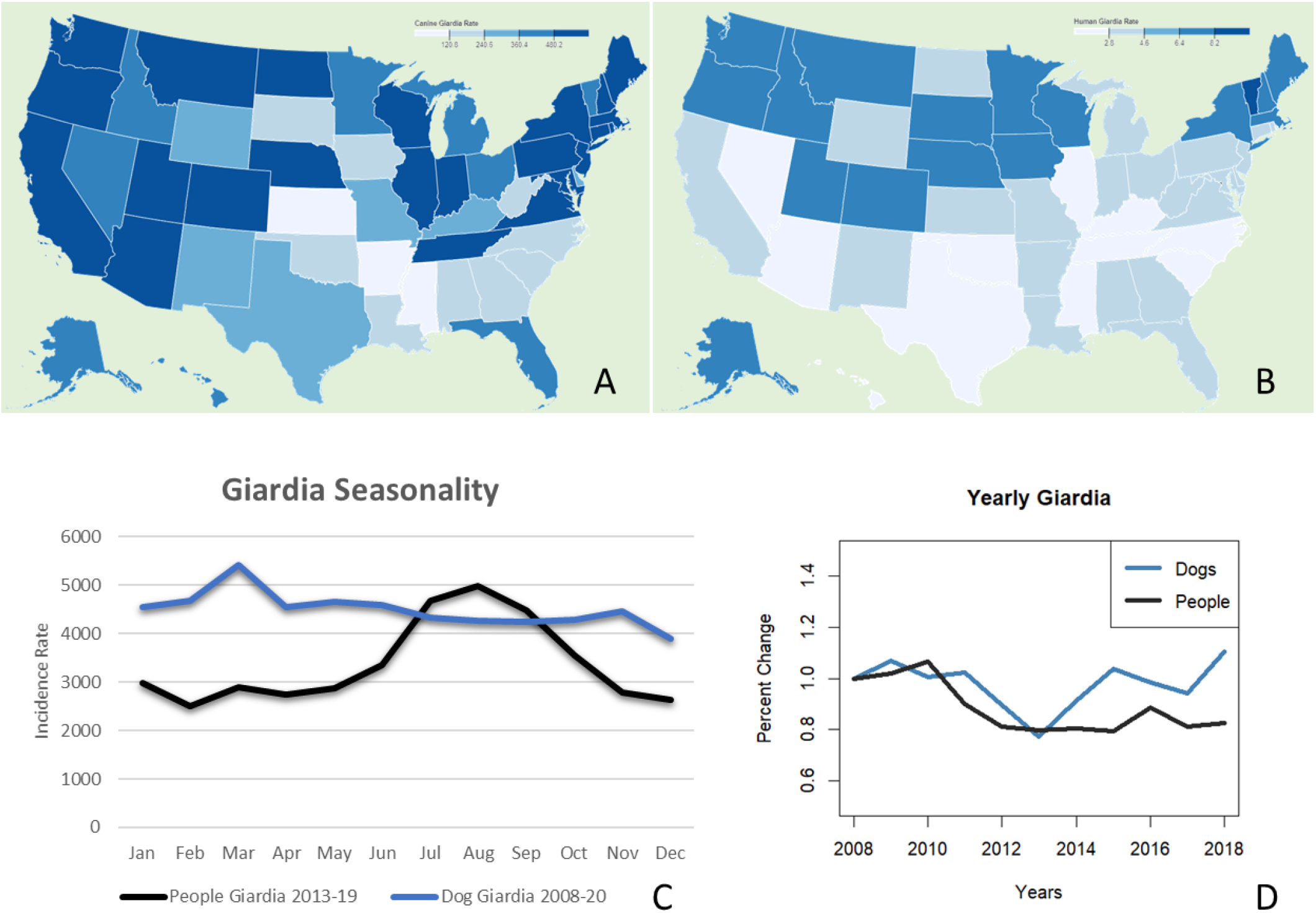
Geographical distribution of Giardia for A) dogs and B) humans – incidence rates. C) Seasonal trends in Giardia in humans and dogs – canine incidence rate per 1000, human incidence rate per 100,000. D) Yearly Giardia trends in humans and dog

The insurance data from dogs show that dogs have a peak incidence in the spring - specifically in March, and the incidence slowly decreases from there. Similarly to how canine Lyme cases have a more spread out seasonality compared to human Lyme cases, canine giardia is much more spread out compared to human cases throughout the year. Humans have a season starting in May, and then peaking in August. The geographic distributions of diagnoses also appear similar during these years – with the northeast and northwest being most impacted and the southeast less impacted. Overall the Giardia incidence rate among insured dogs has been gradually increasing, with a trough year in 2015.

#### Coccidioidomycosis (Valley Fever)

The geographic distribution of Coccidioidomycosis cases in dogs is nearly identical to that in humans, with California and Arizona being the hot-spots within the US. The trend lines for human and canine cases across years appear to be matching each other (see **Figure 4**), with peaks in both species occurring in 2011, and troughs occurring in 2014-2015. Data after 2019 for human cases was not publicly available at the time of analysis.

**Figure 4:**
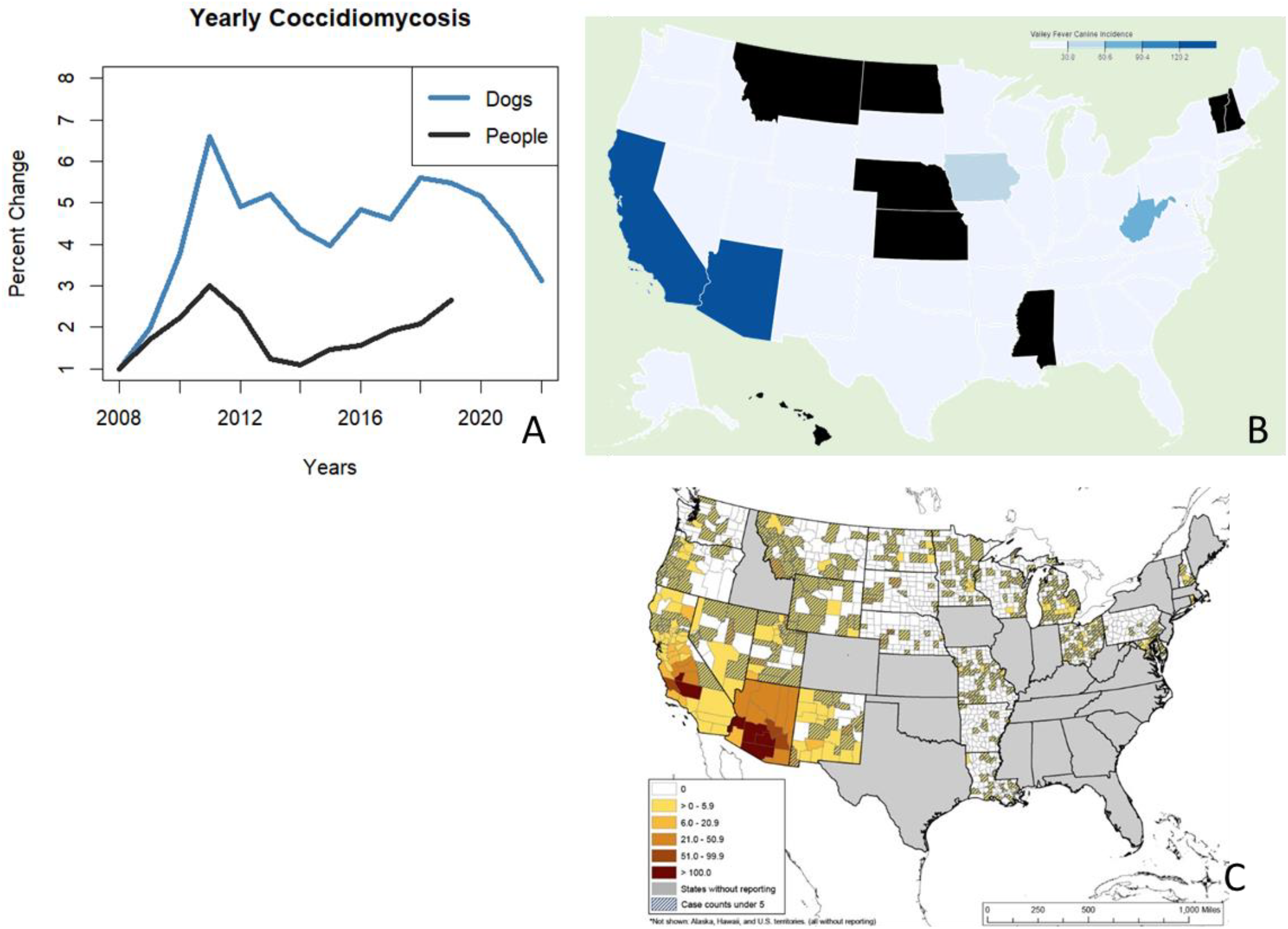
Yearly Coccidioidomycosis trends in dogs and humans b) case counts geographic distribution of coccidioidomycosis – c) county-level human data from CDC.

#### Intestinal Parasites

The intestinal worm incidence rates were highest in New York, New Jersey, South Carolina, Oregon, and Hawaii (see **Figure 5**). Overall the incidence rates have been declining since 2008, with some increases in 2015 and 2020. Diagnoses among dogs are less common in the winter months and more common in the summer, with peak diagnoses occurring in July. The authors also attempted to compare incidence rates for canine intestinal parasites in dogs to the incidence rates for the human conditions caused by these parasites (termed soil-transmitted helminths). Unfortunately the most recently conducted studies of soil-transmitted helminths in the human population were performed in the 1980s, so recent endemic incidence rates for people are not readily available.^19,26^

**Figure 5:**
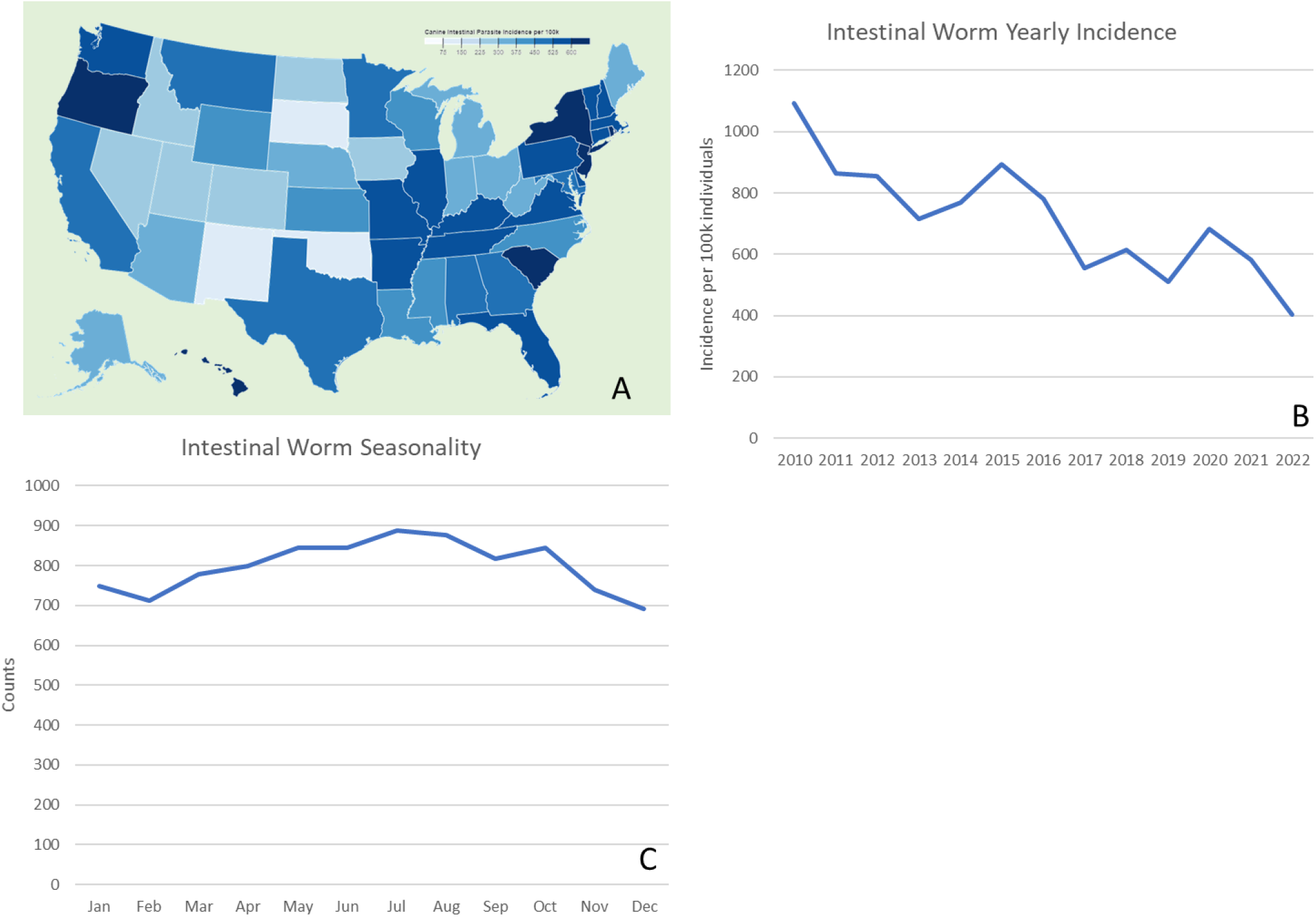
Intestinal worm diagnoses among this population of insured pet dogs: A) geographically B) yearly incidence per 100k, and C) seasonally from 2010-2022

#### Machine Learning

The models showed that the Mean Absolute Error (MAE) for predictions across all the years of data available decreased for each of the models when dog incidence data was added to the model (see **Table 2**). The amount that the MAE is reduced in each case varies, but the addition of dog data improves the model predictions in every case.

**Table 2:**
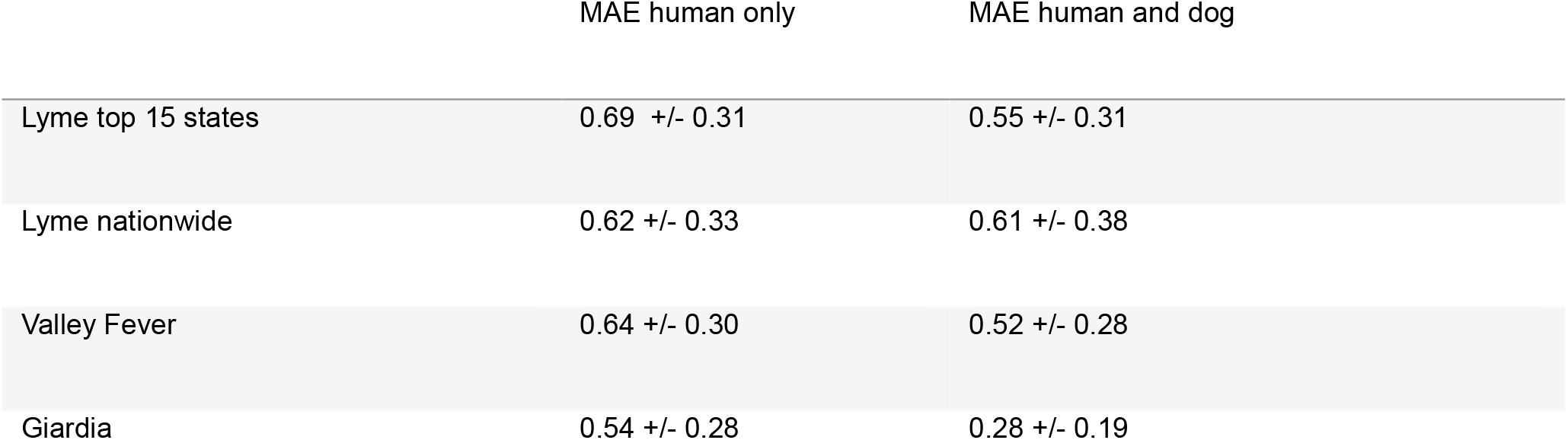
Mean Absolute Error (MAE) +/- standard deviation for the constructed models using human-only incidence information compared to using both human and dog incidence information.

## Discussion

Four very different pathogens were investigated: a vector-borne bacterium, an airborne soil fungus, a water-borne protozoa, and soil-transmitted helminths. The transmission routes in people and dogs are diverse between these diseases, and yet each disease for which human data was available demonstrated a similarity in disease trends between humans and dogs.

Unfortunately, human data on soil-transmitted helminths in the United States is sparse after the 1980s so a direct comparison between canine and human incidence rates was not possible. However, since dogs are the reservoir host for many of the soil-transmitted helminths, active monitoring of intestinal worm diagnoses in dogs would likely assist researchers in the surveillance of these neglected parasitic diseases.

Examining both intestinal pathogens in the canine insurance data, puppies were overwhelmingly the largest group diagnosed with giardia and intestinal parasites, which is consistent with previous studies regarding the prevalence of intestinal parasites in puppies.^27^ The overall differences in lifestages diagnosed with different diseases is helpful to understand which dogs are most susceptible in the epidemiology of these disease groups and can inform both public health and veterinary practitioners. The incidence rate of intestinal worm diagnoses is overall decreasing, which may be due to increased use of monthly preventative medications. The increases in 2015 and 2020 may have been due to momentarily decreased preventative use due to economic factors (e.g. the 2015 stock market selloff, or the COVID-19 pandemic, shutdowns, and decreased access to well care for pets). The rise in diagnoses in summer months may correspond with more pets being outdoors and therefore being more exposed to infection, or also environmental temperatures being conducive for transmission.

With coccidioidomycosis, the peaks and troughs in people and dogs occur in the same years, which is logically consistent with the method of transmission: environmental conditions influence the levels of fungal spores in the air, and when the spores are more present, more dogs and humans are infected by this single agent. Here we successfully showed that incidence rates in both species are temporally similar, suggesting that at least for endemic areas spikes in canine coccidioidomycosis rates should correlate with spikes in human rates during the same years.

Monitoring cases for both species could make the detection of epidemic years more sensitive. It was also demonstrated that amongst this pet insurance dataset, the geographic trends of the disease are identical to the geographical set in humans.

In spite of the difference in giardia assemblages primarily responsible for disease in both people and dogs, the overall incidence rates for both species from 2008-2020 follow similar patterns with peaks and troughs in the trends occurring in similar years. Likely, the conditions that allow for canine giardia to cause a spike in infections are likely the same environmental conditions that allow for human giardia infections to spike within the same year. Conveniently, canine giardia is seasonally highest in spring before human infections begin to rise in May. If it can be demonstrated through future research that spikes in canine giardia in spring precede a spike in human giardia in the same year, active monitoring of canine giardia diagnoses could help public health professionals to implement interventions to protect people with weeks to months of warning. --- It is unclear whether a high Giardia spring season for dogs precedes a high Giardia Summer season for humans, but if this is the case, it is promising again that pet insurance data could be a surveillance tool for human public health.

Of all diseases investigated, Lyme disease trends in the pet insurance data presented the largest possible benefit to public health. Unlike coccidioidomycosis and giardiasis, canine Lyme incidence trends appear to precede human Lyme incidence trends by two years at both the national and top-15 state level. Furthermore, the overall incidence rate in dogs is magnitudes higher than the rate in people, making spikes in the disease within this species easier to detect. If active monitoring of canine Lyme diagnoses can give public health agencies a 2-year lead time on emergences of Lyme, there is the potential that budget planning can earmark funds specifically for Lyme disease interventions. The reason for this 2-year gap in trends is unclear through this analysis: perhaps the 2-year lifecycle of the Ixodes vector plays a role, perhaps the 2-year lag is representative of the length of time required to obtain a diagnosis in people, or some other reason. Regardless of the cause for the 2-year difference, knowing canine Lyme trends can help public health agencies to intervene in advance to improve human health.

The use of pet insurance data in this study is beneficial as it is a large dataset of dogs used to achieve statistically significant results. Insured dogs also have varied breed heritage including purebreds and mixed breeds, live in varied geographic locations, and visit different veterinary practice types (i.e. general practice and referral care facilities). This allows comparison of disease across these variables to paint a cohesive picture for infectious disease modeling and management. For example, this insurance dataset was able to provide comprehensive temporal and geographical data for dog Valley Fever diagnoses without reliance on reference laboratory results and veterinary teaching hospital records, which present additional barriers to data acquisition.

Insured dogs in the data set tended to be puppies and young adults. This is likely due to the lower premium cost for younger dogs and the low probability of a pet having a pre-existing condition. Most pet insurers do not cover medical conditions that occurred prior to the inception of the policy.

There are drawbacks of using pet insurance data as well. Pet owners with pet insurance are able to afford both the premium and the reimbursement payment model and have access to veterinary care. Therefore, opting to use pet insurance data may impact the representation of certain demographics for example, residing in a rural area and retirement status of pet owners may impact whether a pet owner has pet insurance.^28^ This demographic information is important to account for because knowing how and when pet owners seek veterinary care has important implications for the prevention of the spread of infectious disease. For these demographic reasons, perhaps insurance is not the best tool to monitor soil-transmitted helminths in underserved areas.

Additionally, some cases of disease may go unreported because a policyholder did not file a claim after seeking veterinary care. Cases could also be missed if the date of the veterinary invoice occurred prior to the policy inception date, during the policy waiting period, or beyond the 90 day claim submission deadline. In these cases, a condition code may not be assigned. In spite of these drawbacks to pet insurance data, it preliminarily appears to be a very useful tool for monitoring at least three diseases that both humans and dogs share.

The models that were trained with both human and dog incidence information had a lower error score, indicating that across all prediction years, adding dog information to the model improves the predictions overall. These models were very simple, given that there were only a decade worth of data for both humans and dogs, and the yearly incidences were a single data point for each year. It would be valuable to further demonstrate with incidence dates across human and dog data (and not just overall yearly rates) whether the dog data improves the human prediction models for these diseases more granularly – especially given the difference in seasonality between dogs and humans for both Lyme and Giardia.

Fetch pet insurance currently utilizes a machine learning algorithm which predicts disease probabilities based on a pet’s information. This algorithm is individual pet focused at the moment, directed at improving the customer’s experience. However this algorithm could be retooled to be a syndromic surveillance database with population-level predictions for diseases within the pet dog population. If such a tool can accurately model and predict these shared diseases within the dog population, areas of emergence and times of re-emergence could be quickly detected using pet insurance data, and used to alert human health practitioners and public health workers. If active monitoring of pet insurance data can detect, or even predict, bad seasons for canine giardia, Lyme, soil transmitted helminths, or Valley Fever, health departments could be alerted to the issues before human cases begin rising excessively. In the case of Lyme disease, this alert could be several years in advance of when human cases start rising, so public health agencies have the opportunity to earmark funding specifically for tick prevention and Lyme disease awareness education.

## Methods

### Pet insurance data claims codes

The claims used in this study are from dogs insured by Fetch, Inc. from all 50 United States and 12 of 13 Canadian Provinces (Fetch, Inc does not offer pet insurance coverage in Quebec) from years 2008-2022. Fetch maintains a list of disease codes that denotes the medical condition name or clinical signs of disease. Once a policyholder files a claim, a disease code is assigned as stated on the accompanying medical records, invoice, and/or policyholder declaration. The condition codes for each disease included in this study are: Tickborne Diseases (Lyme disease, babesiosis, ehrlichiosis, anaplasmosis, tick bite), Giardiasis (one condition code: Giardia), Fungal Diseases (Coccidioidomycosis, Aspergillosis, general fungal), Intestinal Parasites (ascarids, coccidiosis, intestinal parasites, toxoplasmosis, whipworms, hookworm, roundworm/strongyle, tapeworm, internal parasites). Fetch groups dog ages by lifestages, as defined by the American Animal Hospital Association (AAHA) lifestage guidelines, which accounts for both age and size of dog.^29^

### Pet insurance data analysis

The counts for each disease were tabulated by state/province, year, and month within year. The overall population data was also sorted into these categories to determine the total number of active policies (dog-time). The counts data were normalized and incidence rates were calculated – this was done using a combination of the Excel platform with pivot tables and R studio.

### Human data analysis

Data regarding the yearly trends and monthly trends of Lyme, Giardia, and Coccidioidomycosis (valley fever) were obtained from the CDC website^30–32^– this data is publicly available for download. Giardia data for the years 2013-2019 were available at the time of publication. Lyme data was available for 2008-2021, and Coccidioidomycosis data was available for 1998-2019. The geographic trends for Lyme and Coccidioidomycosis were also obtained from the CDC site. The geographic trends for giardia for the years 2011-2016 were recreated from the choropleth map displayed by Coffey.^33^ The CDC Lyme counts per state were normalized by the 2020 total state population according to the 2020 census data^34^

### Comparing Human data to dog data

Creating direct comparisons of incident rates for every disease group was not possible as the incident rates were not available for the human population. Therefore for seasonal Lyme trends, seasonal Giardia trends, and geographic distribution of coccidioidomycosis were compared based on count data alone. When plotting yearly trends, the incident rates were plotted relative to the 2008 rate for that species. That is, the yearly incident rate was divided by the 2008 incident rate for the species and disease, and those relative measures were plotted on the same indexed chart to compare relative human disease trends and canine disease trends. This method was chosen over the dual-axis chart to emphasize relative trends of the diseases rather than changes to total counts. Specifically for the seasonality of Lyme disease, the counts of dog Lyme diagnoses required a 200x multiplier in order to display the trends lines on the same chart. The human data for Lyme seasonality was available only in counts, so therefore the canine data was also presented in counts for this comparison. In order to display the Giardia incidence rates relative to each other on the same chart, the canine incidence rates are displayed per 1000 dogs, while the human incidence rates are per 100,000 persons.

### Cross-validation Models

The models were created using the R studio package “caret”, and the Mean Absolute Error for the models were calculated using the package “metrics.” Two models were trained for each disease for which both human and dog data were available. The yearly incidence rates for the disease in question were input, and a new variable for the slope of the dog incidence rate were calculated. The columns were normalized using a scaler function. The first model was trained using the previous year’s human incidence information to predict the year’s incidence rate. The second model was trained using the previous year’s human incidence information and the previous year’s dog slope to predict the year’s incidence rate for humans. The notable exception is that for the Lyme data, the two years ago dog slope was used instead of the previous year’s, since the trends were offset by 2 years. The models were trained using a five-fold cross-validation method. The Mean Absolute Error for the cross-validated models was calculated and the two models for each disease were compared.

## Author Contributions

J.O. – writing, data analysis A.M. – writing, review, editing C.D. – data analysis A.R. – writing, review & editing

## Acknowledgements

The authors thank the owners and dogs, and veterinarians for their important contributions.

## Data Availability

Data is provided within the manuscript and all datasets used and/or analyzed during the current study are available from the corresponding author on reasonable request.

